# Multi-nutrients as Adjunctive Treatment for Bipolar Disorder: A randomized-controlled trial

**DOI:** 10.1101/2023.06.23.23291534

**Authors:** Lewis Mehl-Madrona, Patrick McFarlane

**Author notes:** Corresponding author: PO BOX 39, Orono, ME 04473, USA. Disclosure of funding: Funding for this study was made available through a grant from the Foundation for Excellence in Mental Health Care. The authors have declared no conflicts of interest. ClinicalTrials.gov Identifier: NCT03541031. Data Availability: Data is not available for sharing according to our IRB due to the existence of identifying information that is HIPPA protected in the data.

## Abstract

**Introduction:** An open-label trial suggested that a comprehensive micronutrient supplement, Empower Plus Advanced, in combination with Fish Oil, could reduce symptoms in adults with bipolar disorder. A double-blind, randomized, controlled feasibility trial explored the parameters necessary for a large-scale trial.

**Methods:** Participants (N=69) from a family medicine training clinic with diagnoses of bipolar disorder in the electronic health record were randomized in a 3:2 ratio to Multi-nutrients or Placebo. Diagnoses were confirmed via psychiatric interview or chart review (for obvious cases). The primary outcome measure was change on a composite z-score combining changes on the clinical global impressions scale (CGI), changes on the UKU Side Effects Scale, and changes in medication doses. The GLM repeated measures procedure of SPSS compared continuous outcome measures. Chi-square testing compared responders to non-responders.

**Results:** Data were analyzed for 50 participants. The mean difference of the composite z-score for the primary outcome variables was statistically significant (p = 0.019) and for the composite z-score of all variables (primary and secondary) combined (p = 0.047). In non-parametric chisquare analysis, significantly more in the Nutrient group improved on the CGI (rated 1 or 2) (p = 0.04; OR = 4.0; 52% responders vs. 22% in the Placebo Group). All secondary outcome measures showed nonsignificant trends in the expected direction. Patients in both groups made significant improvement in all measures. The only adverse events occurring more among the Multi-nutrient group were nausea and loose stools, not statistically significant.

**Conclusions:** Multi-nutrients show promise for adjunctive treatment of bipolar disorder. We observed substantial benefits for all patients of closer surveillance, medication adjustment (mostly reduction), and increased human contact. Future studies would benefit from use of a longer lead-in period during which medications can be adjusted and participants can decide if they are willing to take Multi-nutrients for an extended time. Our data suggest that primary care patients with bipolar disorder would fare better on lower medications doses and more frequent visits. Further clinical trials are warranted.

## Introduction

Bipolar disorder is a common neuropsychiatric illness with high rates of morbidity and mortality [1]. Despite available medications to treat bipolar disorder, recurrence rates are high [2]. Bipolar disorder is conventionally treated with typical or atypical antipsychotic medications, anti-epileptic medications, and/or lithium. The risks of atypical antipsychotic medications are well described and include the increased risk of acute kidney injury [3], cataracts [4], decreased cognitive function [5], increased risk for myocardial infarction and stroke [6], metabolic syndrome and type 2 diabetes mellitus [7], and dyslipidemia [8]. Related to this, mortality rates are elevated among people with bipolar disorder compared to the general population; men with the diagnosis of bipolar disorder live, on average 13.6 years less than the general population, and for women, 12.1 years less [9].

Anticonvulsant medications can cause encephalopathy and varying degrees of liver damage, even lethal [10]; hypothyroidism [11]; increased fracture risk [12]; mental sluggishness; falls; coordination problems; and aplastic anemia [13].

Lithium is associated with increased risk for acute and chronic kidney injury, hypothyroidism [11], and increased cancer risk [14, 15]. Lithium’s classic “cognitive dulling” effect, with mild impairments in memory, information processing speed, and creativity, has also been empirically confirmed [16].

There are compelling reasons to explore the role of nutrition in the treatment of psychiatric disorders. First, psychiatric disorders are prevalent and costly to families and society [17]. Second, two important studies have cast doubt on the efficacy of the long-term reliance on psychiatric medication [18, 19].

A solid biologic rationale exists for considering improved nutrition for treating mental health problems. A review by Ames and colleagues showed that as many as one-third of known genetic mutations resulted in the corresponding enzyme having an increased Michaelis-Menten coefficient, or Km, which results in decreased binding affinity for a coenzyme, and then a lower rate of reaction [20]. At least 50 human genetic diseases caused by defective enzymes can be remedied or ameliorated by the administration of sufficiently high doses of the vitamin component of the corresponding cofactor, which at least partially compensates for reduced enzymatic activity. For patients who are not able to improve their diet, supplementation with micronutrients may provide equivalent benefit. In fact, Bell et al. showed increased enzyme activity coefficients and improved ratings of depression and cognitive function when geriatric patients diagnosed with depression were supplemented with vitamins B1, B2, and B6 [21].

A recent review showed that the adjunctive use of micronutrients has benefited patients with depression [22]. One hundred years of scientific research has provided promising (though modest) results of using single nutrients to modulate mood swings and irritability [23]. In contrast, research since 2000 on formulas with more than 25 minerals and vitamins (referred to as broad-spectrum) have shown medium size effects [24]. There are now over 20 randomized controlled trials (RCTs) showing that adding micronutrients provides benefit by reducing the impact of stress; reducing anxiety and aggression; and improving mood, irritability, and inattentiveness [25]. One RCT showed that a 25-ingredient micronutrient formula plus some omega-3 fatty acids (EPA and DHA) was associated with a one-third reduction of aggressive offenses in young adult prisoners [26]. In another setting, micronutrients supplementation had a statistically significant impact on emotions of stress related to the 6.3 magnitude earthquake on February 22, 2011, in Christchurch, NZ [27]. These post-disaster results were replicated in a general population following a destructive flood in southern Alberta, Canada [28].

Sarris et al. performed a meta-analysis of studies done of multi-nutrients as adjuncts to psychiatric medication for depression. They found primarily positive results of replicated studies testing *S*-adenosylmethionine (SAMe), methylfolate, omega-3 (primarily EPA or ethyl-EPA), and vitamin D, with positive isolated studies for creatine, folinic acid, and an amino acid combination. They found mixed results for zinc, folic acid, vitamin C, and tryptophan, with nonsignificant results for inositol. They found no major adverse effects aside from minor digestive disturbance. They found a significant and moderate to strong effect in favor of omega-3 fatty acid and a nonsignificant difference from placebo for folic acid. [29].

Another example of an RCT with a broad-spectrum formula was reported in adults with attention deficit hyperactivity disorder (ADHD) [30]. Those consuming the active formula showed greater reductions in symptoms than those taking placebo, with medium-to-large effect sizes. In a subgroup that entered the trial with moderate to severe depression, twice as many people went into remission in the micronutrient group compared to the placebo group. Importantly, the benefits of micronutrients continued through the 1-year follow-up [31]. Rucklidge et al. conducted a blinded, randomized controlled trial of medication-free children with ADHD (7–12 years) assigned to either multi-nutrients or placebo for 10 weeks [32]. Intent-to-treat analyses showed significant between-group differences favoring micronutrient treatment on the Clinical Global Impression (ES = 0.46), with 47% of those on micronutrients identified as ‘much’ to ‘very much’ improved versus 28% on placebo. According to clinicians, 32% of those on micronutrients versus 9% of those on placebo showed a clinically meaningful improvement on inattention, but no group differences on hyperactive-impulsive symptoms (OR = 1.0; 95% CI: 0.4–2.5). Based on clinician, parent, and teacher report, those on micronutrients showed greater improvements in emotional regulation, aggression and general functioning compared to placebo (ES ranged 0.35–0.66).

In an open-label trial that compared 19 patients who were willing to take micronutrients over 24 months to a convenience sample from the same practice, we [33] found reduced doses of medication and reduced number of side effects with equivalent symptom relief among patients diagnosed with psychotic disorders compared to similar patients who were not supplemented. We used EMPowerplus™ (EMP), a broad-spectrum micronutrient formula. The safety and tolerability of EMPowerplus™ has been assessed by Simpson et al., 2011. All clients were evaluated with the Positive and Negative Symptom Scale and the Clinical Global Impression scale at study baseline and after 3, 6, 9, 12, 15, 18, and 24 months. Psychosis was confirmed with clinical interview using DSM IV-TR criteria. All participants had normal physical examinations and laboratory studies. Outcomes were similar for both groups until 15 months, although the micronutrient group used significantly less antipsychotic medication throughout that time (p < 0.001). At 15 months, the micronutrients + medication group began to exhibit fewer symptoms than the medication-only group, a difference that increased at 24 months. We concluded that improved nutrition using micronutrients among people with psychotic disorders allowed them to achieve similar effectiveness at lower doses of medication and fewer side effects than those who didn’t receive supplements.

There are roughly 90 years of scientific literature demonstrating the relevance of dietary nutrients for mental health, forming the rationale for including the use of micronutrients. Some of the earliest research studies on nutrients relevant to mental illness observed irritability and mood problems in people known to be deficient in the B vitamins [34], as well as reporting positive improvements in mental illness when treated with such nutrients as manganese [35, 36] and nicotinic acid [37], regardless of whether or not they were deficient in said nutrients. Although interest in such studies has declined since the introduction of psychiatric medications in the 1950s, recent work on folic acid (vitamin B9) suggests that low levels may be associated with depressive symptomatology and poor response to antidepressant medication [38, 39]. Further discussion follows in our background and significance section at the end of this proposal.

We aimed to examine whether adjunctive micronutrient treatment would permit lower doses of conventional medications to be effective for bipolar disorder with fewer side effects. Many of the side effects of conventional medication are dose-related, so dose reductions can benefit patients in lowering risk for morbidity and mortality. Micronutrients are relatively safe compared to conventional medications and could make a significant difference in the quality of life of patients with bipolar disorder.

## Methods

### Design of Trial

This was a double-blind, randomized controlled trial that intended to assign 120 stable adult outpatients with bipolar disorder, type 1 or 2 (DSM5 criteria) to supplementation with a 36-ingredient vitamin/mineral formula and an omega-3 fatty acid supplement (N=72) or to matched double placebo (N=48) in a 3:2 ratio for a study duration of one year. This ratio was chosen to improve recruitment, expose fewer participants to placebo, and obtain essentially the same power as a 1:1 ratio. One research coordinator who did not have contact with participants was unblinded so she could dispense appropriately for each participant.

### Methods to Minimize Bias

Only one research coordinator having no contact with the subjects was unblinded. All research and clinical staff and all patients were blinded to the study drug. Placebo drug appeared identical to the active micronutrients. The placebos have been manufactured by the same companies that make the actual micronutrients. Low dose riboflavin has been added to the micronutrient placebo to change urine color to mimic active micronutrient dosing. No adverse events led to the Data Safety Monitor breaking the blind.

### Study Site

All recruiting activity occurred at the Center for Family Medicine of Eastern Maine Medical Center.

#### Participants

The population was largely federally funded through Medicare or Medicaid, was largely white or Native American (reflective of the population of Maine), was rural, and was predominantly female (65%).

a. <u>Target Population:</u> Adults (≥ age 18) with a diagnosis of bipolar disorder, type 1 or 2.
b. <u>**Inclusion Criteria.**</u> We enrolled adult outpatients with a diagnosis of bipolar disorder who were receiving care at the Family Medicine Center & Residency Program’s Clinic of Eastern Maine Medical Center in Bangor, Maine. They met the DSM-5 criteria for bipolar disorder (type I or type 2).
c. <u>**Exclusion Criteria**</u> included being medically or psychiatrically unstable (until stabilized), having mineral-related illnesses (none did), having hypervitaminosis, having chronic kidney disease of stage II or higher, being unable to communicate in English or French, and being pregnant. The only exclusions occurred for pregnancy.

We assessed participants monthly and provided them with EMPowerplus^™^ and Wylie’s Finest Alaskan Fish Oil or placebos. We started with 2 capsules twice daily with meals and increased monthly by 2 capsules twice daily to achieve a maximum dose of 8 capsules twice daily at month 4 as this was the dose that we used in previous studies (Mehl-Madrona, et al., 2010; Mehl-Madrona & Mainguy, 2017, 2018). We chose the 2100 mg dose of EPA in the fish oil (three capsules), since 2000 mg was the threshold dose for improving symptoms among people diagnosed with schizophrenia (Peet, et al., 2002). Nagakura, et al. (2000) used 28.6 mg/kg EPA among children with asthma, which corresponds to 1876 mg for a 70 kg adult. The American Heart Association recommends EPA as preventative (Siskovich, et al., 2018) and reviews studies that use as high as 8 gm/day without adverse events. Placebos were enhanced with riboflavin to give a deep yellow appearance to the urine. Olive oil was used for Fish Oil placebo.

### Outcomes

The primary outcome variable was a composite z-score calculated from three separate z-scores for medication dosage (measured in haloperidol equivalents, valproic acid equivalents, lithium dose, sertraline equivalents, or lorazepam equivalents), the CGI score, and the UKU total side effect score [40]. Using the General Linear Modeling Procedure in SPSS for repeated measures, we obtained a comparison of between group differences and within group differences (improvement over time). Covariates were included. Group differences in change scores were tested for statistical significance. The Sidak correction was used for multiple statistical comparisons. Chi-square testing was used to compare responders to non-responders.

Composite z-scores are commonly used when it is important to integrate outcomes that are measured differently into a final result [41]. They have been used, for example, to combine different measures of cognition into a single variable [42] and for measures in which several domains need to be integrated [43].

The secondary endpoints were (1) Number of ED visits for psychiatric reasons and all reasons (2) Number of hospitalizations for psychiatric reasons and all reasons, (3) Scores on the Positive and Negative Symptom Scale, (4) Scores on the Young Mania Scale, (5) Scores on the My Medical Outcomes Profile version 2 (MYMOP-2), (5) Scores on the Hamilton Anxiety Scale, (6) Scores on the Montgomery-Asburg Depression Rating Scale (MADRS), (7) changes in vital signs, including BMI and waist circumference, and (8) Scores on the Mini-Nutritional Assessment Scale.

### Sample size

We used the G*3 Power software from the University of Dusseldorf to calculate sample size for repeated measures analysis of variance and determined that we needed 97 patients for a conservative effect size estimate of 0.4 to obtain 80% power to detect an effect.

We converted doses of antipsychotic medication to haloperidol equivalents using standardized conversion formulas [33] used in previous research [44]. We found the maximum recommended dose for each anticonvulsant from the manufacturers’ web sites and converted patients’ doses to a proportion of the maximal dose, combining portions when patients took more than one anticonvulsant and then converted proportions back to valproic acid dosages. We used the same approach for antidepressants (sertraline equivalents) and benzodiazepines (lorazepam equivalents). The other medications required were sufficiently infrequent as to not requite conversion to another medication. Z-scores were calculated for each class of medication and then combined to obtain a z-score for all medication changes. Changes in the UKU Side Effects Profile were also converted into z-scores. The composite z-score was obtained by adding the three-component z-scores and then dividing by three. We used an intent-to-treat approach for any patient who had available data after the month 1 time point.

## Results

Figure 1 presents the CONSORT flow diagram for the study including numbers in each category. Table 1 tallies the reasons why people who were randomized were excluded from analysis. Table 2 provides the baseline demographic data for our participants. There were no significant differences between the groups for baseline demographics. There were no statistically significant differences in any of our secondary outcome measures between members of the two groups at baseline. Patients averaged a visit to the clinic every 71.6 days, though not necessarily for their mental health.

**Figure 1.**
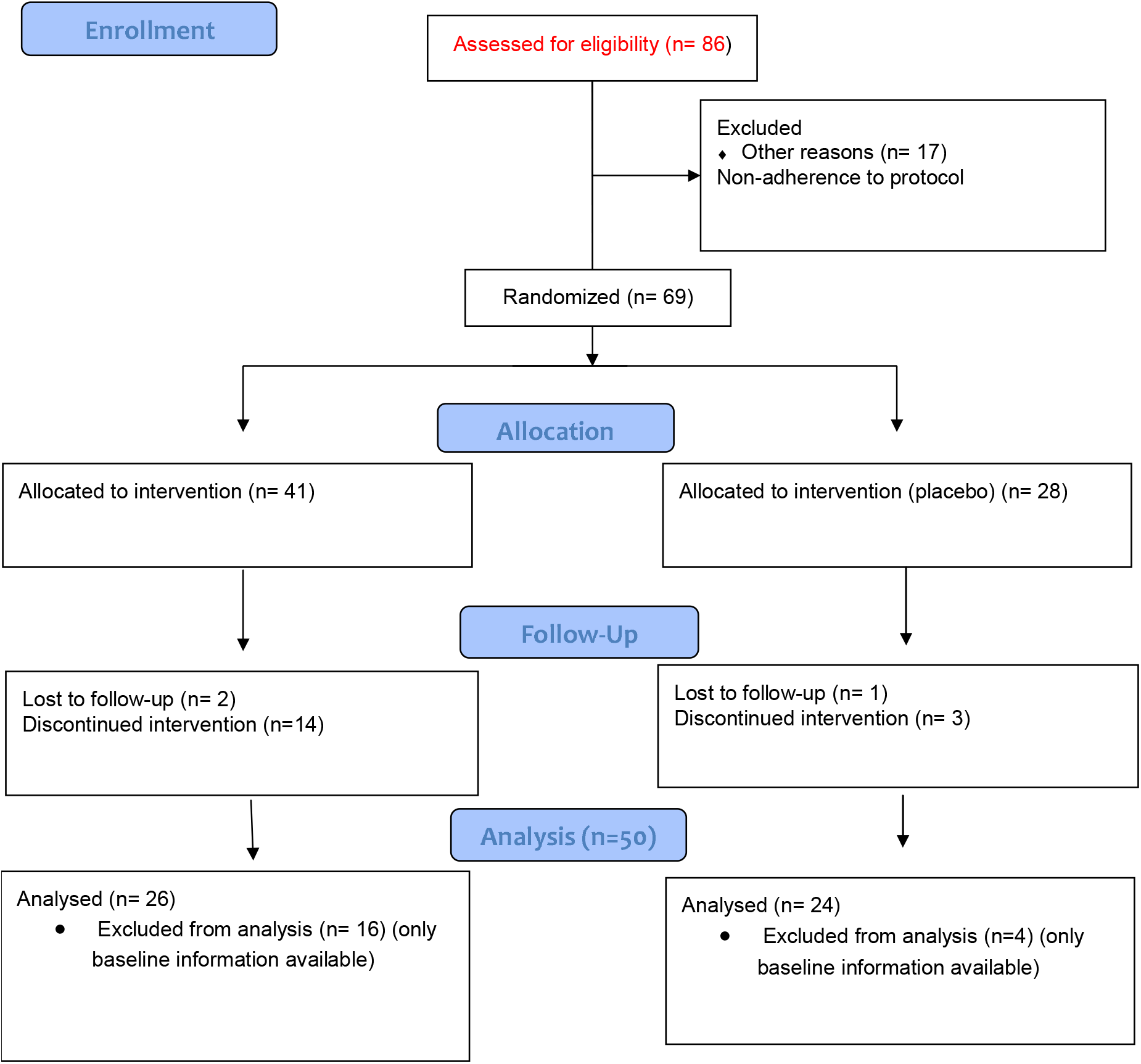
CONSORT Flow Diagram Multi-nutrients.

**Table 1.** Reasons for Discontinuation before the first follow-up appointment.

| Reason for Discontinuation before the first follow-up appointment | Number reporting in the Intervention Group | Number reporting in the Placebo Group |
| --- | --- | --- |
| Moved/left practice/no answer to phone call* | 8 | 1 |
| Rejected possibility of taking placebo | 2 | 0 |
| Enrolled too late in study | 1 | 0 |
| Hives around neck | 1 | 0 |
| Ordered actual vitamins | 2 | 0 |
| Family problems/ too stressed | 1 | 1 |
| Too many pills | 2 | 1 |

\* We can't explain why more people left the practice or moved away in the Intervention Group. It's not logically related to the Multi-Nutrients which would not theoretically cause someone to move or to change medical providers.

**Table 2.** Baseline Demographic Data.

| Table 2. Baseline Demographic Data |  |  |  |  |
| --- | --- | --- | --- | --- |
| Measures at Baseline | Placebo (N= 24) | Intervention (N= 26) | Total (N= 50) | P Value |
| Age (mean ± SD) | 40.61 ± 12.1 | 42.08 ± 13.6 | 41.39 ± 12.853 | 0.694<sup>c</sup> |
| Sex, No. (%) |  |  |  |  |
| Male | 10 (43.5) | 8 (30.8) | 18 (36.7) | 0.390<sup>d</sup> |
| Female | 13 (56.5) | 18 (69.2) | 31 (63.3) |  |
| White race | 23 (100) | 25 (96.2%) | 49 (100%) | 1.000<sup>d</sup> |
| Education<sup>e</sup> No. (%) |  |  |  |  |
| High school or less | 8 (36.4) | 13 (50.0) | 21 (43.8) | 0.393<sup>d</sup> |
| College or graduate | 14 (63.6) | 13 (50.0) | 27 (56.3) |  |
| Marital Status No. (%) |  |  |  |  |
| Married | 5 (20.8) | 8 (30.8) | 13 (26.0) | 0.360<sup>d</sup> |
| Divorced | 10 (41.7) | 6 (23.1) | 16 (32.0) |  |
| Never married | 9 (37.5) | 12 (46.2) | 21 (42.0) |  |
| Social Support No. (%) |  |  |  | 0.131 |
| Single | 20 (83.3) | 18 (69.2) | 38 (76.0) |  |
| Multiple | 4 (16.6) | 8 (30.8) | 12 (24.0) |  |
| Employed or Volunteer in past 30 days No. (%) |  |  |  | 0.433 |
| Yes | 13 (54.1) | 14 (53.8) | 27 (54.0) |  |
| No | 11 (45.8) | 12 (46.2) | 23 (46.0) |  |
| Waist Circ<sup>a</sup>(mean ± SD) | 112.1 ± | 103.42 ± | 107.987 ± | 0.459<sup>c</sup> |
|  | 45.8 | 18.9 | 35.492 |  |
| BMI<sup>b</sup> (mean ± SD) | 33.07 ± 10.8 | 33.51 ± 8.8 | 33.294 ± 9.713 | 0.879<sup>c</sup> |
| BP-Systolic (mean ± SD) | 124.74 ± 18.6 | 121.04 ± 13.3 | 122.78 ± 15.927 | 0.423<sup>c</sup> |
| BP-Diastolic (mean ± SD) | 76.61 ± 9.4 | 74.04 ± 10.1 | 75.24 ± 9.803 | 0.365<sup>c</sup> |
| Heart Rate (mean ± SD) | 80.09 ± 12.9 |  | 78.73 ± 11.028 | 0.425<sup>c</sup> |
| Respiratory Rate (mean ± SD) | 16.87 ± 1.6 |  | 16.37 ± 1.334 | 0.012<sup>c</sup> |

<sup>c</sup> Independent samples *t* test
<sup>d</sup> Fisher's Exact Test

A total of 69 patients were randomized and data were analyzed for 50 patients. The mean difference of the composite z-score for the primary outcome variables favored the Multi-nutrient group and was statistically significant (difference = -0.304; p = 0.019, 95% CI = -0.557 to -0.052). In non-parametric chisquare analysis, significantly more patients in the Multi-Nutrient group improved on the CGI over the course of their participation in the study (p = 0.016; OR = 4.33; 95% CI, 1.269 to 15.490; 53.8% responders vs. 22% in the Placebo Group; see Table 3). For the secondary outcome measurements, the average improvement was not statistically significant between the two groups on any measure but trended toward greater improvement in the Multi-Nutrient group. All patients showed significant improvement over time in all measures. The only adverse events occurring more among the Multi-nutrient group were nausea and loose stools but these were not statistically significant. Mixed linear repeated measures analysis showed a significant interaction of time by group on the Primary Outcome Measure favoring the Multi-Nutrient group (F = 11.404, p = 0.002). None of the other covariates were significant or had significant interactions except for an interaction of the MYMOP Wellbeing Scale with education (F = 5.368; p = 0.003). The blinded research team were unable to accurately guess group assignment.

**Table 3.** Comparisons between Groups.

| Table 3. Comparisons between Groups |  |  |  |
| --- | --- | --- | --- |
| Observation | Placebo | Intervention | p-values |
|  | N= 24 | N=26 |  |
| **Study Visits attended** |  |  |  |
| • Less than or equal to 6 | 13 (54.2%) | 16 (61.5%) | Chi-square = 0.298<br>p = 0.585 |
| • >6 | 11 (45.8%) | 10 (38.5%) |  |
| **Antipsychotic Med Usage** |  |  |  |
| • Decreased | 7 (29.2%) | 4 (15.4%) | Chi-square = 2.743<br>p = 0.098 |
| • Increased | 0 | 3 (11.5%) |  |
| • No Change | 5 (20.8%) | 9 (34.6%) |  |
| • Not on Antipsychotics | 12(50%) | 10 (38.5%) |  |
| **CGI Scores (given monthly; change from baseline to end of study)** |  |  |  |
| • Responders | 5 (20.8%) | 14 (53.8%) | Chi square = 5.773<br>p = 0.016 |
| • Non-Responders | 19 (79.2%) | 12 (46.2%) |  |
| • Worsened over the course of the study (included in non-responders) | 10 (41.7%) | 6 (23.1%) |  |
| **UKU Side Effects Total Scores** |  |  |  |
| • Improved | 18 (75%) | 23 (88.5%) | Chi-square = 1.187<br>p = 0.276 |
| • Worsened | 4 (16.7%) | 3 (11.5%) |  |
| • No Change | 2 (8.3%) | 0 |  |

Table 4 shows the changes in time in the CGI from baseline to time 5 (which is 12 months). Average improvement from baseline to six months (time 3) was statistically significant for all participants (time 3). Improvement leveled at six months and remained statistically significantly better than baseline throughout. Substantial improvement occurred in all measures over time with most of the improvement completed by six months (hence, the lack of significance of the “change in the last month” on the CGI. Two measures approached significance – the CGI Severity of Illness rating and the reduction in neurological side effects.

**Table 4.** Differences in outcome measures over time for the entire sample. Minus signs indicate improvement.

|  | Baseline Value | Change from Baseline to Time 5 | Standard Error | Significance | 95% Confidence interval |
| --- | --- | --- | --- | --- | --- |
| CGI: Severity of Illness | 3.82 ± 0.92 | -1.131 | 0.147 | < 0.001 | 0.836 to 1.427 |
| CGI: Average Interval Change during the past month | 3.62 ± 1.16 | -0.417 | 0.220 | 0.064 | -0.025 to 0.859 |
| CGI: Change from Worst Phase of Illness | 2.80 ± 1.03 | -0.720 | 0.178 | <0.001 | 0.361 to 1.078 |
| MYMOP Symptom 1 | 4.48 ± 1.25 | -3.921 | 0.182 | <0.001 | 3.555 to 4.288 |
| Basis-24 Score | 61.12 ± 9.09 | -4.620 | 1.249 | <0.001 | 2.100 to 7.140 |
| MADRS | 18.3 ± 8.61 | -6.886 | 1.344 | <0.001 | 4.1555 to 9.576 |
| Hamilton Anxiety | 16.38 ± 11.84 | -6.131 | 1.274 | <0.001 | 3.562 to 8.700 |
| Young Mania | 19.45 ± 4.6 | -2.835 | 0.850 | 0.002 | 1.126 to 4.544 |
| Total UKU Side Effects | 19.4 ± 6.18 | -2.616 | 0.713 | <0.001 | 1.180 to 4.052 |
| UKU Neurological Side Effects | 10.19 ± 2.10 | -1.833 | 0.650 | 0.007 | 0.524 to 3.141 |
| UKU Psychological Side Effects | 13.14 ± 6.66 | -4.936 | 0.907 | <0.001 | -6.759 to -3.113 |
| UKU Autonomic Side Effects | 5.18 ± 4.35 | -2.715 | 0.685 | <0.001 | 1.337 to 4.093 |
| UKU Side Effects – Other Symptoms | 5.01 ± 2.10 | -1.810 | 0.240 | 0.008 | 0.477 to 3.112 |

Table 5 shows the comparisons of the differences in outcome measures between the two groups (a minus sign favors the Multinutrient group). Despite the small sample size, statistically significant differences were seen in the composite outcome score for the three primary outcome measures combined and all outcome measures combined.

**Table 5.** Comparisons of differences in outcome measures between the two groups (a minus sign favors the Multi-nutrient group).

| Variable | Difference between groups | Standard Error | Significance | 95% Confidence Interval |
| --- | --- | --- | --- | --- |
| CGI: Severity of Illness | -0.413 | 0.235 | 0.085 | -0.886 to 0.059 |
| CGI: Change in the last month | -0.301 | 0.240 | 0.215 | -0.784 to 0.181 |
| CGI: Change from the Worst Phase of Illness | -0.093 | 0.145 | 0.525 | -0.199 to 0.365 |
| MYMOP Symptom 1 | -0.115 | 0.142 | 0.422 | -0.402 to 0.171 |
| Basis-24 Score | -2.090 | 2.757 | 0.424 | -7.501 to 3.214 |
| MADRS | -2.143 | 2.663 | 0.425 | -3.691 to 3.171 |
| Hamilton Anxiety | -1.274 | 2.754 | 0.646 | -4.264 to 6.812 |
| Young Mania | -1.135 | 1.891 | 0.551 | -4.936 to **2.667** |
| Total UKU Side Effects |  |  |  |  |
| UKU Neurological Side Effects | -1.739 | 0.892 | 0.058 | -0.059 to 3.536 |
| UKU Psychological Side Effects | -0.593 | 1.926 | 0.759 | -4.465 to 3.279 |
| UKU Autonomic Side Effects | -1.157 | 1.231 | 0.352 | -3.632 to 1.318 |
| UKU Side Effects – Other Symptoms | -1.021 | 1.282 | 0.201 | -2.117 to 2.390 |
| Composite z-score Primary Outcome Measures | -0.304 | 0.125 | 0.019<br>(t = -2.429) | -0.557 to -0.052 |
| Composite z-score Secondary Outcome Measures | -0.135 | 0.146 | 0.362<br>(t = -0.922) | -0.429 to 0.160 |
| Composite z-score all measures combined | -0.232 | 0.113 | 0.047<br>(t = -2.403) | -0.461 to -0.003 |

Table 6 shows changes in medication use over time. The reduction in medications for all participants was statistically significant over time for antipsychotics, anticonvulsants, and antidepressants. The only hospitalization occurred in a participant who discovered her husband having sex with another woman and went to the Emergency Department to request an overnight observation admission as a preventive measure. She stayed for one night and was in the placebo group. No emergency department visits were related to multi-nutrient use and no significant differences were noted in number of E.D. visits between groups. The cost per completed subjects averaged $7102;00. Thirty-seven percent of those costs were spent on direct research and 63% of those costs were spent on regulatory matters related to the FDA and to our IRB. All referrals to our study came from family doctors or psychotherapists. No referrals came from psychiatrists or psychiatric nurse practitioners, and some actively discouraged their patients from participating in the study.

**Table 6.** Changes in medication use over time.

| Drug | Multinutrient Group, Beginning Dose | Ending Dose | Placebo Group, Beginning Dose | , Ending Dose | Significance |
| --- | --- | --- | --- | --- | --- |
| Antipsychotics (chlorpromazine equivalents) | 13 people, 334.8 mg | 13 people, 151.0 mg | 15 people, 307.6 mg | 15 people, 196.0 mg | p = 0.17 |
| Anticonvulsants (Valproate mg equivalents) | 17 people, 786.7 mg | 17 people, 631.2 mg | 20 people, 525.7 mg | 20 people, 497.9 mg | p = 0.073 |
| Antidepressants (Sertraline mg equivalents) | 14 people, 160.8 mg | 14 people, 154.3 mg | 20 people, 207.9 mg | 20 people, 150.3 mg | p = 0.254 |
| Lithium | 4 people, 720 mg | 4 people, 720 mg | 3 people, 750 mg | 3 people, 750 mg | NS |
| Hydroxyzine | 5 people, 79 mg | 5 people, 59 mg | 5 people, 115 mg | 5 people, 73 mg | NS |
| Opiates in morphine equivalents | 5 people, 63 mg | 5 people, 68 mg | 8 people, 70 mg | 8 people, 72 mg | NS |
| Psychostimulants in methylphenidate equivalents | 4 people, 61.9 mg | 4 people, 69.4 mg | 6 people, 50.3 mg | 6 people, 38.8 mg | NS |
| Requip | 1 person, 1 mg | 1 person, 0 mg | 1 person, 3 mg | 1 person, 3 mg | NS |
| Baclofen | 5 people, 51 mg | 5 people, 25 mg | 2 people, 25 mg | 2 people, 10 mg | NS |
| Buspirone | 2 people, 37 mg | 2 people, 10 mg | 2 people, 15 mg | 2 people, 15 mg | NS |
| Benzodiazepines in clonazepam equivalents | 3 people, 8.6 mg | 3 people, 8.6 mg | 2 people, 3 mg | 2 people, 0.75 mg | NS |
| Clonidine | 4 people, 0.14 mg | 4 people, 0.11 mg | 4 people, 0.23 mg | 4 people, 0.23 mg | NS |

## Conclusions

Despite the small sample size of this feasibility trial, we found statistically significant improvements in the primary outcome variable in favor of Multi-nutrients. The odds ratio of receiving benefit from Multi-nutrients was 4.33. This strongly suggests that Multi-nutrients as adjuncts or primary therapies should be pursued in future studies.

Given our high rate of dropouts, we recommend a lead-in period of 3-4 months to determine if participants are willing to take the micronutrients over an extended period and to stabilize their medications. Significant reductions in medication dosages occurred over the course of the study along with reductions in side effects with improvements in all the outcome measures. Patients were seen more often by being in the study (every month) compared to approximately every 2 months and not necessarily for mental health. Improvements plateaued by 6 months for all participants. Thus, a six-month trial would be reasonable instead of our 12 months. The effects of medication optimization and increased contact, which happened during our study, may have led to substantial benefit that overshadowed the effects of the multi-nutrients.

Simply being in the study was associated with statistically significantly improvement in all measures. This suggests that increased frequency of visits could improve the symptoms of patients diagnosed with bipolar disorder. The patients in this setting were symptomatic at a level below needing hospitalization, but still distressed. More severely symptomatic patients might increase the power of future studies, though we note that no psychiatrists in our region made referrals to the study, and some actively discouraged their patients from participating. We found that it was more difficult than we anticipated to retain subjects, presumably related to the number of pills that people needed to take. The cost per subject was also greater than we anticipated, primarily related to the increased regulatory requirements imposed by our Institutional Review Board (IRB). Since we conducted this study in a Primary Care setting, every Primary Care visit was seen as an Adverse Event, including falls on ice, insect bites, and strains and sprains. This produced many Adverse Events to report to the IRB. We also recommend that future studies use a means of quantifying subject intake of Multi-nutrients through an assay of at least one component.

Our primary care patients led complicated lives. They struggled with the social determinants of health, including lack of income, lack of employment, and lack of transportation. Another strategy would be to recruit only stable patients who do not struggle with these issues and then determine if their medication doses could be reduced when micronutrients are added. That might require multiple recruitment centers.

Sarris et al. conducted the most similar study to ours -- a multi-site, 8-week, double-blind, randomized, controlled trial involving 158 outpatients with a diagnosis of major depressive disorder [45]. Their attrition rate was high as was ours. Their intervention consisted of a nutraceutical combination of S-adenosyl methionine, Folinic acid, Omega-3 fatty acids, 5-HTP, Zinc picolinate, and relevant co-factors versus placebo. Placebo was nominally superior to the nutraceutical combination in reducing depression scores on the MADRS with a non-significant Group x Time interaction. Response rates were 40% for the active intervention and 51% for the placebo; remission rates were 34% and 43% for active and placebo groups, respectively. They found no significant differences between groups on any other secondary depression, anxiety, psychosocial, or sleep outcome measures. No significant differences occurred between groups for total adverse effects, aside from more nausea in the active group. They concluded that a “very high placebo response rates suggest [that] a placebo run-in design may have been valuable. Our results suggest the same – that a run-in period of about 4 months is necessary. Sarris, et al., however, did not use a comprehensive formula of multi-nutrients and did not follow participants long enough to see effects.

The substantial concerns of our IRB for adverse effects of multi-nutrients were not met. Reducing medication doses while adding micronutrients and fish oil did not appear to harm anyone. In bipolar disorder one expects that some patients’ symptoms will worsen over time. Fewer worsened in the multi-nutrient group on clinician observation and the averages were not significantly different between groups on any outcome measures. Our participants did better on all outcome measures with lower doses of medication and fewer side effects. Reduced intensity of surveillance and fewer outcome measures could also reduce the costs of future studies.

## Data Availability

Data contains protected health information and according to our IRB cannot be shared.

## Acknowledgements

The authors wish to thank our funders and colleagues Dr. Raymond Baxter, Dr. Jessica Bloom-Foster, Julie Weidman, Cindy Whited, Denise Michaud, Krysta Anderson, Jessica Kneser, Jennifer Trumbo, Evelyn Preston, Karianne Sjostedt, Andrew Pritchard, Nicole Cournoyer, and Tina Temples, CCRC at EMMC Clinical Research Center for their support and assistance. We are also grateful to True Hope, Inc., which provided us with micronutrients and placebo at no cost and to Wylie’s Alaska’s Finest Fish Oil, who provided us with fish oil and placebo at no cost.

## References

1. Walker, E.R., R.E. McGee, and B.G. Druss, Mortality in mental disorders and global disease burden implications: a systematic review and meta-analysis. JAMA psychiatry, 2015. 72(4): p. 334–341.

2. Gitlin, M.J. and D.J. Miklowitz, The difficult lives of individuals with bipolar disorder: A review of functional outcomes and their implications for treatment. Journal of Affective Disorders, 2017. 209(February): p. 147–154.

3. Ryan, P.B., et al., Atypical Antipsychotics and the Risks of Acute Kidney Injury and Related Outcomes Among Older Adults: A Replication Analysis and an Evaluation of Adapted Confounding Control Strategies. Drugs & Aging, 2017: p. 1–9.

4. Chu, C.-S., et al., Association between antipsychotic drug use and cataracts in patients with bipolar disorder: A population-based, nested case-control study. Journal of Affective Disorders, 2017. 209: p. 86–92.

5. Franza, F., et al., Risk and efficacy in cognitive functions in Bd Ii with atypical antipsychotics: a two-year open study. Bipolar Disorders, 2016. 18: p. 135.

6. Prieto, M.L., et al., Long-term risk of myocardial infarction and stroke in bipolar I disorder: A population-based Cohort Study. Journal of affective disorders, 2016. 194: p. 120–127.

7. Samaras, K., Cardiometabolic Risk and Monitoring in Psychiatric Disorders, in Cardiovascular Diseases and Depression. 2016, Springer. p. 305–331.

8. Rajagopalan, K., et al., Adherence to Lurasidone and other Atypical Antipsychotics among Patients with Bipolar Disorder: A Real World Assessment. Journal of Health & Medical Economics, 2016.

9. Laursen, T.M., Life expectancy among persons with schizophrenia or bipolar affective disorder. Schizophrenia research, 2011. 131(1): p. 101–104.

10. Patel, N., et al., Reversible Encephalopathy due to Valproic Acid Induced Hyperammonemia in a Patient with Bipolar I Disorder: A Cautionary Report. Psychopharmacology Bulletin, 2017. 47(1): p. 40.

11. Lambert, C.G., et al., Hypothyroidism risk compared among nine common bipolar disorder therapies in a large US cohort. Bipolar disorders, 2016. 18(3): p. 247–260.

12. Shen, C., et al., Association between use of antiepileptic drugs and fracture risk: a systematic review and meta-analysis. Bone, 2014. 64: p. 246–253.

13. Hogan, C.S. and M.P. Freeman, Adverse Effects in the Pharmacologic Management of Bipolar Disorder During Pregnancy. Psychiatric Clinics of North America, 2016. 39(3): p. 465–475.

14. Huang, R.-Y., et al., Use of lithium and cancer risk in patients with bipolar disorder: population-based cohort study. The British Journal of Psychiatry, 2016: p. bjp. bp. 116.181362.

15. Correll, C.U., et al., Effects of antipsychotics, antidepressants and mood stabilizers on risk for physical diseases in people with schizophrenia, depression and bipolar disorder. World Psychiatry, 2015. 14(2): p. 119–136.

16. Koek, R.J., Tips, cautions, and successes in treating bipolar patients with lithium. Psychiatric Times, 2015. 32(4): p. 52–52.

17. Chong, H.Y., et al., Global economic burden of schizophrenia: a systematic review. Neuropsychiatric disease and treatment, 2016. 12: p. 357.

18. Harrow, M., T. Jobe, and R. Faull, Does treatment of schizophrenia with antipsychotic medications eliminate or reduce psychosis? A 20-year multi-follow-up study. Psychological Medicine, 2014. 44(14): p. 3007–3016.

19. Wunderink, L., et al., Recovery in remitted first-episode psychosis at 7 years of follow-up of an early dose reduction/discontinuation or maintenance treatment strategy: long-term follow-up of a 2-year randomized clinical trial. JAMA psychiatry, 2013. 70(9): p. 913–920.

20. Ames, B.N., I. Elson-Schwab, and E.A. Silver, High-dose vitamin therapy stimulates variant enzymes with decreased coenzyme binding affinity (increased Km): relevance to genetic disease and polymorphisms. The American journal of clinical nutrition, 2002. 75(4): p. 616–658.

21. Bell, I.R., J.S. Edman, and F.D. Morrow, Vitamin B1, B2, and B6 augmentation of tricyclic antidepressant treatment in geriatric depression with cognitive dysfunction. Journal of The American College of Nutrition, 1992. 11(2): p. 159–163.

22. Sarris, J., et al., Nutritional medicine as mainstream in psychiatry. The Lancet Psychiatry, 2015. 2(3): p. 271–274.

23. Kaplan, B.J., et al., Vitamins, minerals, and mood. Psychological bulletin, 2007. 133(5): p. 747.

24. Popper, C.W., Single-micronutrient and broad-spectrum micronutrient approaches for treating mood disorders in youth and adults. Child and adolescent psychiatric clinics of North America, 2014. 23(3): p. 591–672.

25. Rucklidge, J.J. and B.J. Kaplan, Broad-spectrum micronutrient formulas for the treatment of psychiatric symptoms: a systematic review. Expert Review of Neurotherapeutics, 2013. 13: p. 49–73.

26. Gesch, C.B., et al., Influence of supplementary vitamins, minerals and essential fatty acids on the antisocial behaviour of young adult prisoners Randomised, placebocontrolled trial. The British Journal of Psychiatry, 2002. 181(1): p. 22–28.

27. Rucklidge, J.J., et al., Shaken but unstirred? Effects of micronutrients on stress and trauma after an earthquake: RCT evidence comparing formulas and doses. Human Psychopharmacology: Clinical and Experimental, 2012. 27(5): p. 440–454.

28. Kaplan, B.J., et al., A randomised trial of nutrient supplements to minimise psychological stress after a natural disaster. Psychiatry research, 2015. 228(3): p. 373–379.

29. Sarris, J., et al., Adjunctive nutraceuticals for depression: a systematic review and metaanalyses. American Journal of Psychiatry, 2016. 173(6): p. 575–587.

30. Gordon, H.A., et al., Clinically significant symptom reduction in children with attentiondeficit/hyperactivity disorder treated with micronutrients: an open-label reversal design study. Journal of child and adolescent psychopharmacology, 2015. 25(10): p. 783–798.

31. Rucklidge, J.J., et al., Psychiatric comorbidities in a New Zealand sample of adults with ADHD. Journal of attention disorders, 2014: p. 1087054714529457.

32. Rucklidge, J.J., et al., Vitamin-mineral treatment improves aggression and emotional regulation in children with ADHD: a fully blinded, randomized, placebo-controlled trial. Journal of child psychology and psychiatry, 2018. 59(3): p. 232–246.

33. Mehl-Madrona, L. and B. Mainguy, Adjunctive Treatment of Psychotic Disorders with Micronutrients. The Journal of Alternative and Complementary Medicine, 2017.

34. Hoobler, B.R., Symptomatology of Vitamin B deficiency in infant. Journal of the American Medical Association, 1928. 91: p. 307–310.

35. English, W.M., Report of the treatment with manganese chloride of 181 cases of schizophrenia, 33 of manic depression, and 16 other defects of psychoses at the Ontario Hospital, Brockville, Ontario. American Journal of Psychiatry, 1929. 9: p. 569–580.

36. Reed, G.E., Use of manganese chloride in dementia praecox. Canadian Medical Association Journal, 1929. 21: p. 46–49.

37. Sydenstricker, V.P. and H.M. Cleckley, The effect of nicotinic acid in stupor, lethargy and various other psychiatric disorders. American Journal of Psychiatry, 1941. 98: p. 83–92.

38. Alpert, J.E., et al., Nutrition and depression: Focus on folate. Nutrition, 200. 16: p. 544–546.

39. Fava, M., et al., Folate, vitamin B12, and homocysteine in major depressive disorder. Am J Psychiatry, 1997. 154(3): p. 426–8.

40. Lindström, E., et al., Patient-rated versus clinician-rated side effects of drug treatment in schizophrenia. Clinical validation of a self-rating version of the UKU Side Effect Rating Scale (UKU-SERS-Pat). Nordic Journal of Psychiatry, 2001. 55(sup44): p. 5–69.

41. Andrade, C., Z Scores, Standard Scores, and Composite Test Scores Explained. Indian journal of psychological medicine, 2021. 43(6): p. 555–557.

42. Lim, Y.Y., et al., Sensitivity of composite scores to amyloid burden in preclinical Alzheimer’s disease: Introducing the Z-scores of Attention, Verbal fluency, and Episodic memory for Nondemented older adults composite score. Alzheimer’s & Dementia: Diagnosis, Assessment & Disease Monitoring, 2016. 2: p. 19–26.

43. Song, M.-K., et al., Composite variables: when and how. Nursing research, 2013. 62(1): p. 45.

44. Andreasen, N.C., et al., Antipsychotic dose equivalents and dose-years: a standardized method for comparing exposure to different drugs. Biological psychiatry, 2010. 67(3): p. 255–262.

45. Sarris, J., et al., Nutraceuticals for major depressive disorder-more is not merrier: An 8-week double-blind, randomised, controlled trial. Journal of Affective Disorders, 2019. 245: p. 1007–1015.

